# Physical activity, respiratory physiotherapy practices, and nutrition among people with primary ciliary dyskinesia in Switzerland

**DOI:** 10.1101/2022.05.11.22274957

**Authors:** Yin Ting Lam, Eva SL Pedersen, Leonie D Schreck, Leonie Hüsler, Helena Koppe, Fabiën N Belle, Christian Clarenbach, Philipp Latzin, Swiss PCD research group, Claudia E Kuehni, Myrofora Goutaki

**Affiliations:** Institute of Social and Preventive Medicine, University of Bern, Bern, Switzerland; Graduate School for Cellular and Biomedical Sciences, University of Bern, Bern, Switzerland; Center for Primary Care and Public Health (Unisanté), University of Lausanne, Lausanne, Switzerland; Department of Pulmonology, University Hospital Zurich, Zurich, Switzerland; Paediatric Respiratory Medicine, Children’s University Hospital of Bern, University of Bern, Bern, Switzerland

**Keywords:** Primary ciliary dyskinesia, epidemiology, orphan disease, physical activity, physiotherapy, nutrition

## Abstract

**Aims of the study:** We know little about the level of physical activity, respiratory physiotherapy practices, and nutritional status of people with primary ciliary dyskinesia (PCD), although these are important aspects of patients with chronic respiratory disease. We assessed physical activity, respiratory physiotherapy practices, and nutritional status among people with PCD in Switzerland, investigated how these vary by age, and identified factors associated with regular physical activity.

**Methods:** We sent a postal questionnaire survey to people with PCD enrolled in the Swiss PCD registry (CH-PCD), based on the standardised FOLLOW-PCD patient questionnaire. We collected information about physical activity, physiotherapy, respiratory symptoms, and nutritional status. We calculated the metabolic equivalent (MET) to better reflect the intensity of the reported physical activities. To assess nutritional status, we extracted information from CH-PCD and calculated participants’ body mass index (BMI).

**Results:** Of the 86 questionnaires we sent, 74 (86% response rate) were returned from 24 children and 50 adults. The median age at survey completion was 23 years [IQR (interquartile range) 15–51], and 51% were female. Among all 74 participants, 48 (65%) performed sports regularly. Children were vigorously active (median MET 9.1; IQR 7.9–9.6) and adults were moderately active (median MET 5.5; IQR 4.3-6.9). 59 participants (80%) reported performing some type of respiratory physiotherapy. However, only 30% of adults saw a professional physiotherapist compared with 75% of children. Half of the participants had normal BMI; one child (4%) and two adults (4%) were underweight. People who were regularly physically active reported seeing a physiotherapist more often.

**Conclusions:** Our study is the first to provide patient-reported data about physical activity, respiratory physiotherapy, and nutrition among people with PCD. Our results highlight that professional respiratory physiotherapy, exercise recommendations, and nutritional advice are often not implemented in the care of people with PCD in Switzerland. Multidisciplinary care in specialised centres by teams including physiotherapists and nutrition consultants could improve the quality of life of people with PCD.

## Introduction

Due to increased respiratory workload, chronic respiratory diseases affect exercise capacity and nutritional status. These diseases are often complicated by recurrent infections that contribute further to increased metabolic demand [1-4]. Promoting a good level of physical activity and monitoring nutritional status are crucial for maintaining respiratory health; thus, they are important parts of managing chronic lung diseases. In the chronic lung disease primary ciliary dyskinesia (PCD), genetic mutations cause structural and functional changes of motile cilia, which reduce mucociliary clearance [5]. Chronic respiratory symptoms and recurrent infections of the upper and lower airways are common in PCD [6]. Therefore, among people with PCD physical activity is recommended for improving airway clearance, in addition to specific respiratory physiotherapy practices [7-9]. We know from other chronic lung diseases, such as cystic fibrosis (CF), that physical activity is associated with improved sputum expectoration. Particularly beneficial are higher intensity activities followed by respiratory physiotherapy [10,11].

Yet, we know little about patients with PCD and their levels of physical activity, respiratory physiotherapy practices, and nutritional status. So far, the few studies involving physical exercise among people with PCD focused on muscle strength measurements and aerobic cardiorespiratory performance [12-15]. No studies examined the level of physical activity of people with PCD in their everyday lives. Regarding nutritional status, no studies assessed appetite and nutritional advice given to people with PCD. A large study from the international PCD (iPCD) cohort reported that children younger than 9 years had lower body mass index (BMI) z-scores than healthy peers [16]. A single centre study of 43 children in the United Kingdom studied nutrition using impedance spectroscopy [17]. Both studies suggested patients with PCD should receive nutritional advice to improve growth and delay lung disease progression [17,18]. In Switzerland where there is no centralised care for people with PCD, we have no information about supportive care. Our study describes physical activity, respiratory physiotherapy practices, and nutritional status among people with PCD in Switzerland, investigates how these vary by age and identifies factors associated with regular physical activity.

## Methods

### Study design and population

Our national cross-sectional questionnaire survey was nested in the Swiss PCD registry (CH-PCD). CH-PCD is a patient registry (www.clinicaltrials.gov; identifier NCT03606200) enrolling people with confirmed or clinical diagnoses of PCD in Switzerland [19]. Patients with a clinical PCD diagnosis, have a strong clinical suspicion e.g. situs anomalies, persistent cough, persistent rhinitis, chronic or recurrent upper or lower respiratory infections and history of neonatal respiratory symptoms as term infants but have not completed the diagnostic algorithm and have negative or ambiguous results for the tests performed so far [19-21].

Our reporting conforms with the Strengthening the Reporting of Observational studies in Epidemiology (STROBE) statement [22]. Detailed information about the study design is published elsewhere [23]. CH-PCD received ethical approval from the Cantonal Ethics Committee of Bern in 2015 (KEK-BE: 060/2015). We obtained written informed consent from participants or parents of participants younger than 14 years.

In February 2020, we sent a postal questionnaire to all people with PCD enrolled in the CH-PCD. After we mailed the questionnaire by post, we distributed the questionnaire at the PCD outpatient clinic in Bern to an additional five people who newly enrolled in CH-PCD (figure 1). After 2–3 weeks, we sent a reminder by post to everyone who had not yet returned questionnaires.

**Figure 1:**
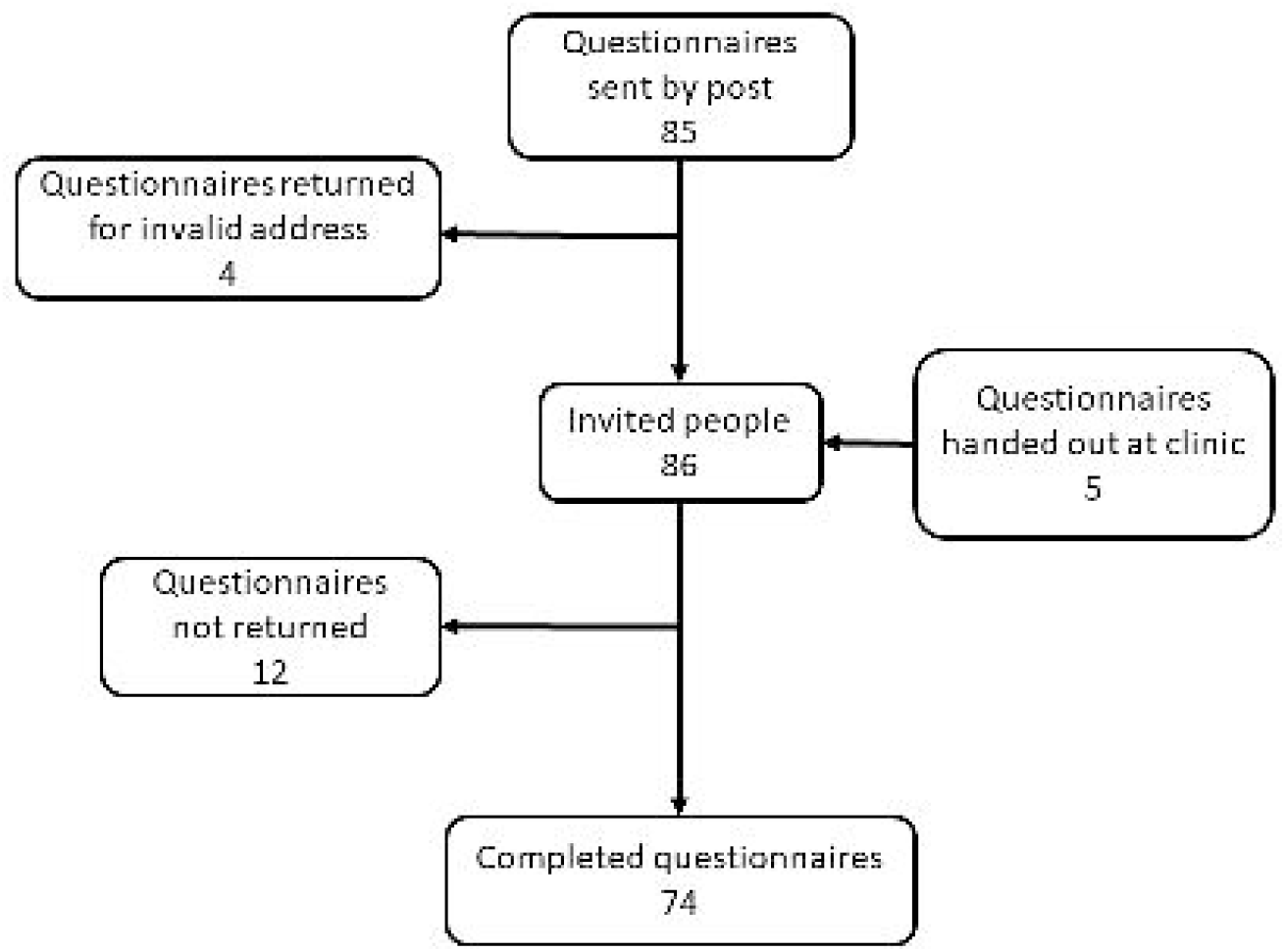
Flowchart of people with PCD living in Switzerland and their parents who were invited and participated in the survey.

### Questionnaire

The questionnaire was based on the FOLLOW-PCD questionnaire (version 1.0), a PCD-specific standardised questionnaire, and part of a follow-up form developed by an international PCD expert group [24]. The FOLLOW-PCD questionnaire’s main domains included chronic respiratory symptoms from the past three months and health-related behaviours, such as physical activity during the past 12 months [23]. Our study used all original questions from the FOLLOW-PCD questionnaire and included additional questions about respiratory physiotherapy and nutritional intake. We added a section on respiratory physiotherapy based on the relevant clinical module from the FOLLOW-PCD form [24]. We reviewed the physiotherapy-related questions with respiratory physiotherapists in Switzerland to ensure they would be understandable by participants and relevant to local techniques. Lastly, we added more questions about nutritional advice in Switzerland based on expert suggestions.

We developed new questions in German, then a native French speaker with knowledge about PCD translated them into French. Questionnaires were age-specific for adults (ages ≥18 years), adolescents (ages 14–17 years), and parents of children (ages 0–13 years) with PCD available in German and French [23].

### Physical activity

We asked if participants performed sports, what type of sports, and how many hours per week. For school-age children, we asked if they played school or any additional sports. We classified physical activity into five categories: aerobic activity, such as endurance or cardio activity; muscle strengthening; bone strengthening; balance activity; and flexibility activity [25]. If multiple types of physical activity were listed, we reported them all.

Since types of physical activity differ in intensity of cardiorespiratory exertion, we used metabolic equivalents (METs) to better reflect the intensity of reported activities. The MET of the adult compendium is described as the ratio of work metabolic rate to the resting metabolic rate [26]. One MET is defined as the amount of oxygen consumed sitting at rest; it is equivalent to 3.5 ml oxygen/kg body weight/min. We calculated the total MET per week based on participants’ activities. Since there is no standardised reference, we added a MET combination of gymnastics (3.8 MET) and children’s games (5.8 MET) to the total MET per week for children who always performed school sports. For children attending school sports sometimes or usually, we added one-third or two-thirds of the MET combination of gymnastics and children’s games, respectively. We categorised physical activity in three MET intensity categories: light (<3.0 METs), moderate (3.0–5.9 METs), and vigorous (≥6.0 METs) [27].

### Nutrition

We assessed participant nutritional status by collecting information about appetite, physician advice for increasing caloric intake during the past 12 months, and calculating participant BMIs. If increasing caloric intake was recommended, we asked about specific recommendations, such as increases of energy-dense food, larger portion size, or meal frequency. Furthermore, we assessed if recommendations had been successful increasing energy and if body weight stabilized or increased. Other questions revolved around intake of oral nutritional supplements.

To calculate BMI, we used height and weight data from the CH-PCD measured in the hospital or private practices closest to the time when participants completed the survey. We calculated BMI by dividing weight in kilograms by height in meters squared (kg/m^2^). We classified BMI for adults as underweight (<18.5 kg/m^2^), normal (≥18.5 to <25), overweight (≥25.0 to <30), or obese (≥30.0 to <35) by World Health Organisation (WHO) standards (WHO) [28]. For children and adolescents aged 5–17 years, we calculated sex and age-specific BMI z-scores based on the 2007 WHO references [29]. We defined thinness (<-2 z-scores), normal (−2 to 1 z-scores), overweight (1 to 2 z-scores), and obesity (>2 z-scores).

### Respiratory physiotherapy

We collected information on respiratory physiotherapy practices during the past three months, including if participants performed any physiotherapy; whether they had seen a professional physiotherapist; and which respiratory practices they used for upper and lower airway clearance, such as nose blowing, nasal rinsing, specific airway clearance techniques, and use of breathing exercise aids. We asked about the use of inhalation for the upper and lower airways, particularly with isotonic (0.9% sodium chloride) or hypertonic (>0.9% sodium chloride) saline. Lastly, we asked how difficult airway clearance was for them during this period and if they felt any improvement of their respiratory symptoms after respiratory physiotherapy.

### Respiratory symptoms

The questionnaire asked about several upper and lower respiratory symptoms and their frequency during the past three months. We defined symptoms as frequent if they were reported daily or often. Reported frequency of all symptoms ranged from daily, often, sometimes, and rarely to never [23].

### Possible factors associated with regular physical activity

We categorised physical activity into regular (once a week or more) and irregular (less than once a week). We considered the following factors as possibly associated with regular physical activity: experiencing respiratory symptoms, such as chronic nasal symptoms, cough, shortness of breath, and sputum production, that might influence physical activity performance; performing any physiotherapy; and seeing professional physiotherapists.

### Statistical analysis

We described characteristics of the population, physical activity, respiratory physiotherapy practices, and nutritional status in the total population and separately among children (<18 years) and adults with PCD. For continuous variables, we used median and interquartile range (IQR); for categorical variables, we used numbers and proportions. We used Wilcoxon rank-sum and Pearson’s Chi square to study differences between children and adults and factors possibly associated with regular physical activity. We calculated Wilson 95% confidence intervals (CI) for proportions. We performed all analyses with Stata version 16 (StataCorp LLC, Texas, USA).

## Results

We invited 86 people; 74 (86%) participants returned the questionnaire (figure 1). The median age at survey completion was 23 years (IQR 15–51) and 38 (51%) were female. Among participants, 24 were children or adolescents (referred as children from now onwards) of whom 17 attended school. Of the 50 adults, 42% were employed full-time (80–100%), while 18% were retired or on disability pension (table 1). In the past 12 months, most children and adults (64%) missed fewer than one week of school or work due to PCD-related symptoms; few adults (10%) missed more than 2 weeks. About half of participants (46%) lived in areas with little or no traffic. Age and sex did not differ between questionnaire respondents and non-respondents (data upon request). Missing responses to individual survey questions were less than 5%.

**Table 1:** Characteristics of people with PCD in Switzerland participating in the survey, overall and by age group (N=74)

|  | **Total N (%)** | **Children <18 y N (%)** | **Adults ≥18 y N (%)** | **p-value** |
| --- | --- | --- | --- | --- |
| **Number of participants** | 74 (100) | 24 (100) | 50 (100) |  |
| Age, median (IQR) | 23 (15–51) | 11 (6–15) | 32 (23–57) |  |
| Sex, female | 38 (51) | 7 (29) | 31 (62) |  |
| **Occupation** |  |  |  |  |
| Student/Preschool | 29 (39) | 24 (100) | 5 (10) |  |
| 81–100% employment | 21 (28) | - | 21 (42) |  |
| 61–80% employment | 2 (3) | - | 2 (4) |  |
| 41–60% employment | 5 (7) | - | 5 (10) |  |
| ≤40% employment | 8 (11) | - | 8 (16) |  |
| Retired/disability pension | 9 (12) | - | 9 (18) |  |
| **Missing (pre)school or work days due to PCD in the past 12 months** |  |  |  | 0.345 |
| 0–6 days | 46 (62) | 15 (63) | 31 (63) |  |
| 1–2 weeks | 8 (11) | 4 (17) | 4 (8) |  |
| >2 weeks | 5 (7) | 0 (0) | 5 (10) |  |
| Not in school yet/retired | 12 (16) | 3 (12) | 9 (18) |  |
| Not reported | 3 (4) | 2 (8) | 1 (1) |  |
| **Area of residence** |  |  |  | 0.949 |
| Street with dense traffic | 17 (23) | 6 (25) | 11 (22) |  |
| Street with moderate traffic | 23 (31) | 7 (29) | 16 (32) |  |
| Street with little or no traffic | 34 (46) | 11 (46) | 23 (46) |  |

PCD: Primary ciliary dyskinesia. y: years. All characteristics are presented as N and column % with the exception of age presented as median and IQR: interquartile range.

### Physical activity

Among all 74 participants, 48 (65%) performed sports regularly (table 2). All 17 schoolchildren attended school sports; 14 of them (82%) participated always. The seven children not yet in school, were all physically active. Categories of sports did not differ by age. The most common type of physical activity were aerobic activities, e.g. children played soccer or hockey (25%), whereas adults preferred jogging or walking (56%). We found children were vigorously active (median MET 9.1; IQR 7.9–9.6) and adults were moderately active (median MET 5.5; IQR 4.3–6.9). Adults spent more time (median 7 hours/week; IQR 3.5–12) performing light sports when compared with children who spent most of their time (median 4 hours/week; IQR 3–7) performing vigorous sports. We present the most reported physical activities in supplemental table S1.

**Table 2:** Physical activity of people with PCD in Switzerland, overall and by age group (N=74)

|  | **Total N (%)** | **Children <18 y N (%)** | **Adults ≥18 y N (%)** | **p-value** |
| --- | --- | --- | --- | --- |
| **Number of participants** | 74 (100) | 24 (100) | 50 (100) |  |
| **Regular sports<sup>#</sup>** | 48 (65) | 13 (54) | 35 (70) | 0.182 |
| **School sports attendance** |  | 17 (71) | NA |  |
| Always |  | 14 (59) | NA |  |
| Usually |  | 2 (8) | NA |  |
| Sometimes |  | 1 (4) | NA |  |
| Not yet in school |  | 7 (29) | NA |  |
| **Sports categories** |  |  |  |  |
| Aerobic activity | 37 (50) | 9 (38) | 28 (56) | 0.136 |
| Muscle strengthening | 21 (28) | 5 (21) | 16 (32) | 0.319 |
| Bone strengthening | 4 (5) | 2 (8) | 2 (4) | 0.440 |
| Balance | 5 (7) | 2 (8) | 3 (6) | 0.708 |
| Flexibility | 3 (4) | 1 (4) | 2 (4) | 0.973 |
| **Total MET per week<sup>§</sup>** | 6.8 (4.8–8.9) | 9.1 (7.9–9.6) | 5.5 (4.3–6.9) | 0.000 |
| **Sports hours/week by MET categories<sup>&</sup>** |  |  |  |  |
| Light (<3.0 METs) | 7 (3.5–12) | 0 (0) | 7 (3.5–12) |  |
| Moderate (3.0–5.9 METs) | 3 (2–3) | 2.5 (2–6) | 3 (2–3) | 0.336 |
| Vigorous (≥6.0 METs) | 4 (3–6) | 4 (3–7) | 5 (3–5.5) | 0.198 |

PCD: Primary ciliary dyskinesia. y: years. NA: not applicable.
Characteristics are presented as N and column %, metabolic equivalent (MET=1 kcal/kg/hour) described in median and IQR: interquartile range.
<sup>#</sup> For children, this refers to additional sports besides school sports. <sup>§</sup> Average sports intensity per week in MET (median, IQR) including school sports for schoolchildren <sup>&</sup> Reported hours per week participants spent performing sports by MET categories (median and IQR).

### Respiratory physiotherapy

Overall, most participants (59; 80%) reported they performed some type of respiratory physiotherapy; of them, 18 children (75%) and 28 adults (56%) daily. Only 15 adults (30%) saw a professional physiotherapist in comparison to 18 children (75%) (*p*=0.008). Regarding upper airway physiotherapy, 69% of the participants blew their noses; most daily (table 3). Over half of participants (61%) performed nasal rinsing; eight children (33%) and 13 adults (26%) daily. Only 21 participants (28%) performed inhalations for the upper airways; eight children (33%) and five adults (10%) daily. Among all participants, 11 (17%) used saline for upper airway inhalation, nine of them hypertonic (>0.9%) saline. We found no differences in upper airway physiotherapy practices between children and adults apart from nose blowing and nasal rinsing, which were more common in children.

**Table 3:** Upper airway physiotherapy practices of people with PCD in Switzerland, overall and by age group (N=74)

|  | **Total N (%)** | **Children <18 y N (%)** | **Adults ≥18 y N (%)** | **p-value** |
| --- | --- | --- | --- | --- |
| **Number of participants** | 74 (100) | 24 (100) | 50 (100) |  |
| **Nose blowing** |  |  |  | 0.021 |
| Yes, daily | 37 (50) | 10 (42) | 27 (54) |  |
| Yes, often | 8 (11) | 5 (21) | 3 (6) |  |
| Yes, sometimes | 3 (4) | 2 (8) | 1 (2) |  |
| Yes, rarely | 0 (0) | 0 (0) | 0 (0) |  |
| Yes, only during colds | 3 (4) | 3 (12) | 0 (0) |  |
| No/not reported | 23 (31) | 4 (17) | 19 (38) |  |
| **Nasal rinsing** |  |  |  | 0.021 |
| Yes, daily | 21 (28) | 8 (33) | 13 (26) |  |
| Yes, often | 11 (15) | 4 (17) | 7 (14) |  |
| Yes, sometimes | 6 (8) | 5 (21) | 1 (2) |  |
| Yes, rarely | 3 (4) | 0 (0) | 3 (6) |  |
| Yes, only during colds | 4 (6) | 4 (17) | 0 (0) |  |
| No/ not reported | 29 (39) | 3 (12) | 26 (52) |  |
| **Inhalations for the upper airways** |  |  |  | 0.146 |
| Yes, daily | 13 (18) | 8 (33) | 5 (10) |  |
| Yes, often | 4 (5) | 1 (4) | 3 (6) |  |
| Yes, sometimes | 3 (4) | 0 (0) | 3 (6) |  |
| Yes, rarely | 1 (1) | 0 (0) | 1 (2) |  |
| Yes, only during colds | 0 (0) | 0 (0) | 0 (0) |  |
| No/ not reported | 53 (72) | 15 (63) | 38 (76) |  |
| **Upper airways inhalation with saline** | 11 (15) | 7 (29) | 4 (8) | 0.237 |
| - isotonic (0.9% sodium chloride) | 2 (3) | 2 (8) | 0 (0) |  |
| - hypertonic (>0.9% sodium chloride) | 9 (12) | 5 (21) | 4 (8) |  |
| **Upper airways inhalation with other medication** | 5 (7) | 3 (13) | 2 (4) |  |

PCD: Primary ciliary dyskinesia. All characteristics are presented as N and column %. y: years.

Regarding lower airway physiotherapy, 49 participants (67%) applied some airway clearance technique, such as autogenic drainage; 21 participants (28%) daily (table 4). The most common aid used for airway clearance was a flutter (19%) or positive expiratory pressure combined with flutter (7%). Overall, 65% performed inhalations for lower airways; 63% of children and 42% of adults daily, mainly with hypertonic saline. 19 participants (26%) reported difficulty clearing their lower airways; 13 children (54%) and 18 adults (36%) reported their lower respiratory symptoms improved after physiotherapy. Lower airway physiotherapy practices did not differ between children and adults.

**Table 4:** Lower airway physiotherapy practices of people with PCD in Switzerland, overall and by age group (N=74)

|  | Total N (%) | Children <18 y N (%) | Adults ≥18 y N (%) | p-value |
| --- | --- | --- | --- | --- |
| **Number of participants** | 74 (100) | 24 (100) | 50 (100) |  |
| **Airway clearance techniques<sup>#</sup>** |  |  |  | 0.735 |
| Yes, daily | 21 (28) | 8 (33) | 13 (26) |  |
| Yes, often | 8 (11) | 2 (8) | 6 (12) |  |
| Yes, sometimes | 13 (18) | 4 (17) | 9 (18) |  |
| Yes, rarely | 5 (7) | 3 (13) | 2 (4) |  |
| Yes, only during colds | 2 (3) | 1 (4) | 1 (2) |  |
| No/ not reported | 25 (33) | 6 (25) | 19 (38) |  |
| **Use of aid for airway clearance** |  |  |  | 0.565 |
| Flutter | 14 (19) | 5 (21) | 9 (18) |  |
| PEP | 3 (4) | 2 (8) | 1 (2) |  |
| Flutter and PEP | 5 (7) | 2 (8) | 3 (6) |  |
| Blow in straw/nozzle | 3 (4) | 3 (13) | 0 (0) |  |
| Vibrating device | 2 (3) | 2 (8) | 0 (0) |  |
| No/ not reported | 47 (63) | 10 (42) | 37 (74) |  |
| **Inhalation for the lower airways** |  |  |  | 0.233 |
| Daily | 36 (49) | 15 (63) | 21 (42) |  |
| Often | 4 (5) | 1 (4) | 3 (6) |  |
| Sometimes | 6 (8) | 0 (0) | 6 (12) |  |
| Only during colds | 2 (3) | 1 (4) | 1 (2) |  |
| No/ not reported | 26 (35) | 7 (29) | 19 (38) |  |
| **Lower airways inhalation with saline** |  |  |  | 0.515 |
| Yes | 35 (47) | 14 (58) | 21 (42) |  |
| isotonic (0.9% sodium chloride) | 4 (5) | 1 (4) | 3 (6) |  |
| hypertonic (>0.9% sodium chloride) | 31 (42) | 13 (54) | 18 (36) |  |
| **Lower airways inhalation with other medication** | 28 (38) | 9 (38) | 19 (38) |  |
| **Difficulty of clearing lower airways** |  |  |  | 0.581 |
| Easy | 42 (57) | 12 (50) | 30 (60) |  |
| Difficult | 19 (26) | 8 (33) | 11 (22) |  |
| I don't know/ not reported | 13 (17) | 4 (17) | 9 (18) |  |

PCD: Primary ciliary dyskinesia. All characteristics are presented as N and column %. y: years. PEP: positive expiratory pressure. <sup>#</sup> Airway clearance techniques e.g. autogenic drainage

### Nutrition

Weight and height information from clinical visits near survey completion (within 12 months, usually a few months apart) was available for 53 (72%) participants: 16 children and 37 adults. Median BMI was 22.3 kg/m2 (IQR 20.4–25.0) for adults and median BMI z-score was 1.0 (IQR 1.0– 1.0) for children (table 5). Half of participants had normal BMI; only one child (4%) and two adults (4%) were underweight/thin. There was no difference between children and adults in BMI categories. Overall, seven people reported decreased appetite (9%). Physicians recommended increased caloric intake to three children (13%) and three adults (6%); three people (4%) ate larger meal portions, two increased their meal frequency, and one child received energy-dense food. Five adult participants reported ingesting hypercaloric drinks; four only during periods of increased physical activity or colds. Increased caloric intake and hypercaloric drinks helped four patients stabilise or gain weight. One child and two adults reported these measures increased their energy.

**Table 5:** Nutritional information of people with PCD in Switzerland, overall and by age group (N=74)

|  | **Total N (%)** | **Children <18 y N (%)** | **Adults ≥18 y N (%)** | **p-value** |
| --- | --- | --- | --- | --- |
| **Number of participants** | 74 (100) | 24 (100) | 50 (100) |  |
| **BMI median (IQR)** |  |  | 22.3 (20.4–25.0) |  |
| **BMI z-score median (IQR)** |  | 1.0 (1.0–1.0) |  |  |
| **BMI categories** |  |  |  | 0.894 |
| Thinness/underweight | 3 (4) | 1 (4) | 2 (4) |  |
| Normal weight | 37 (50) | 12 (50) | 25 (50) |  |
| Overweight | 10 (14) | 2 (8) | 8 (16) |  |
| Obesity | 3 (4) | 1 (4) | 2 (4) |  |
| Missing | 21 (28) | 8 (34) | 13 (26) |  |
| **Physician recommendation to increase caloric intake by** |  |  |  |  |
| Energy-dense food | 1 (1) | 1 (4) | 0 (0) |  |
| Larger meal portions | 3 (4) | 1 (4) | 2 (4) |  |
| Increased meal frequency | 2 (3) | 1 (4) | 1 (2) |  |
| **Use of hypercaloric drinks in the past 12 months** |  |  |  |  |
| Yes | 5 (7) | 0 (0) | 5 (10) |  |
| **Weight gained/stabilised due to increased caloric intake** |  |  |  |  |
| Yes | 4 (5) | 1 (4) | 3 (6) |  |

PCD: Primary ciliary dyskinesia. y: years. BMI: body mass index. NA: not applicable. All characteristics are presented as N and column % with the exception of BMI and BMI z-score presented as median and IQR: interquartile range.
BMI was available for 16 children and 37 adults. BMI categories were based on the World Health Organisation (WHO) 2007 standards, using BMI in kg/m<sup>2</sup> for adults (underweight <18.5, normal weight ≥18.5 to <25, overweight ≥25.0 to <30, obesity ≥30.0 to <35), and calculating BMI z-scores based on the WHO references for children (thinness <-2 z-scores, normal weight -2–1 z-scores, overweight 1–2 z-scores, obesity >2 z-scores).

### Factors associated with regular physical activity

We found no difference in frequency of reported nasal symptoms, cough, shortness of breath, and sputum production between participants who were regularly or less physically active (figure 2). Respiratory physiotherapy was not associated with regular physical activity. However, regularly physically active people were less likely to see professional physiotherapists (*p*=0.008).

**Figure 2:**
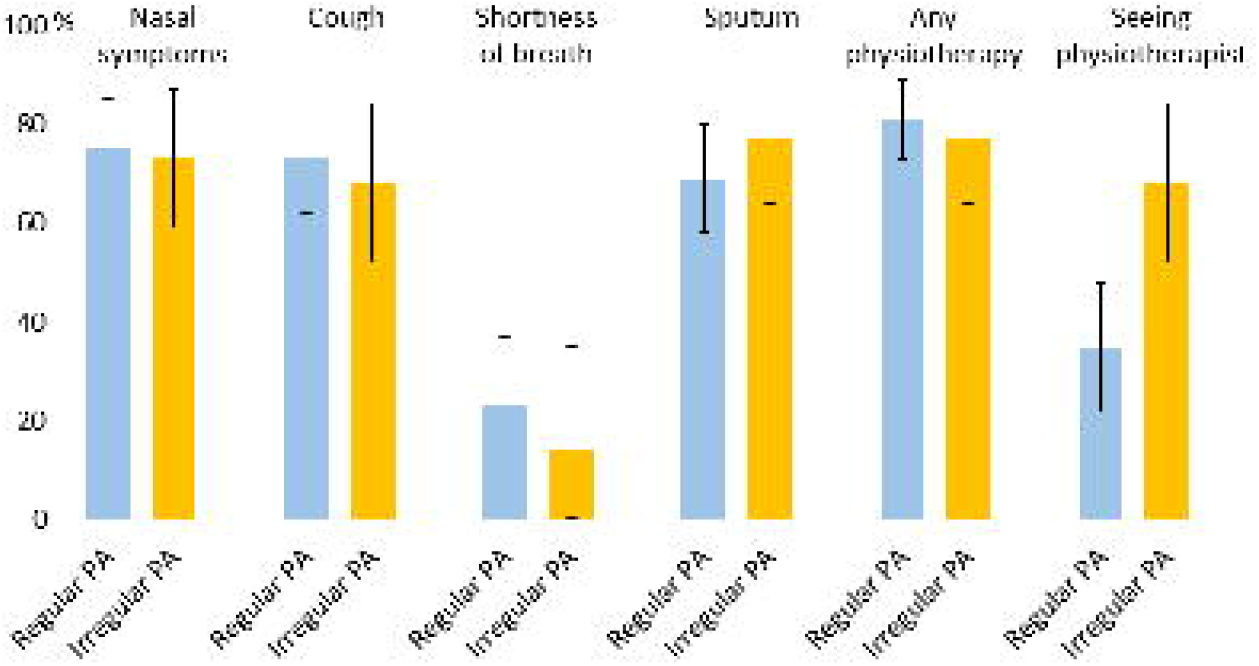
Frequency of reported respiratory symptoms and use of respiratory physiotherapy in the past three months in people with PCD in Switzerland who perform regular (at least once a week) physical activity (PA) compared to people who are less physically active.

## Discussion

Our study is the first to provide patient-reported data on physical activity, respiratory physiotherapy, and nutrition for people with PCD. Our results suggest people with PCD in Switzerland are health-oriented, physically active, and normal weight; they also perform respiratory physiotherapy.

A strength of our study includes our questionnaire. We based our survey on the PCD-specific standardised FOLLOW-PCD questionnaire [24], allowing for future comparisons with larger multicentre studies. We also developed additional questions based on input from experts, such as physiotherapists. Further, we nested our study in the national PCD registry, which provided objectively measured BMI data. Other strengths include a remarkable response rate of 86% and a low proportion of missing survey data.

Our study has several limitations. Although we invited all people enrolled in CH-PCD, selection bias is possible because PCD in Switzerland is underdiagnosed [19]. The COVID-19 pandemic could have affected physical activity. However, since more than 75% of the questionnaires were returned by March 2020 before lockdown in Switzerland started, we expect any effect to be small. Furthermore, most participants reported outdoor activities, which were not affected by COVID-19 restrictions. As many questions referred to the last 12 months, there is a risk of recall bias. We expect answers reflected mostly health-related behaviours closer to date of survey completion. Since FOLLOW-PCD does not include validated physical activity questions, we calculated MET for reported activities to describe exercise intensity. So far, MET is standardised for adults only; however, to describe and compare the intensity of physical activity for the whole study population, we calculated it for all participants.

Previous studies among people with PCD showed reduced physical fitness in hospital settings. For example, a recent single-centre study of 27 Turkish children with PCD reported reduced muscle strength and endurance [15]. Another single-centre study with 22 Danish children with PCD found reduced cardiopulmonary fitness [14]. Children in our study reported a median moderate to vigorous activity of 6.5 hours/week, which is lower than the average 7.5 hours/week among 10–14 year-old children and 6.5 hours/week among 15–19 year-old adolescents who perform additional sports besides attending school sports in Switzerland [30]. However, it is comparable to the level of recreational sports (median 3.0 hours/week, IQR 1–5) of children who survived cancer in Switzerland [31]. In a multicentre service evaluation of over 300 children with PCD in England, all children were trained and advised to perform regular airway clearance physiotherapy [32]. In our population, most people reported doing respiratory physiotherapy but only 18 children (75%) and 28 adults (56%) on a daily basis. Despite having chronic respiratory symptoms, most people with PCD in Switzerland have normal BMI, which is lower than the national average of three people with overweight, and one person with obesity of every ten adults in Switzerland [33]. Our study included very few children younger than age 9 so it was not possible to see whether this group had a lower BMI z-score as in the large iPCD cohort study [18].

Most participants were physically active, despite reporting frequent respiratory symptoms. This might indicate that people with PCD are accustomed to their chronic symptoms and exercise despite them. Or they use physical activity as a method for airway clearance to relieve symptoms, explaining why fewer people who exercised regularly saw a professional physiotherapist. People might replace respiratory physiotherapy with physical activity. Exercise is a recommended method of airway clearance for patients with CF. However, it remains unclear whether it can replace physiotherapy or to what extent for patients with CF or PCD [8,34]. Although physical activity aids in airway clearance of larger airways, peripheral airway dysfunction persists with mucus obstruction [35].

Our study is the first to provide patient-reported data on health-related behaviours of people diagnosed with PCD. Our results highlight the importance of implementing exercise recommendations, professional respiratory physiotherapy, and nutritional advice that are currently missing in the routine care of people with PCD in Switzerland. Multidisciplinary care in specialised centres by teams including physiotherapists and nutrition consultants could improve the daily lives of people with PCD in Switzerland.

## Data Availability

Data may be shared upon reasonable request.

## Author contribution

M Goutaki and CE Kuehni developed the concept and designed the survey. YT Lam, M Goutaki, and L Hüsler organised the survey, then cleaned and standardised the data. YT Lam performed the statistical analyses supervised by M Goutaki and E Pedersen. YT Lam, M Goutaki, and E Pedersen drafted the manuscript. All authors commented and revised the manuscript. YT Lam and M Goutaki take final responsibility for the content.

## Acknowledgments

We thank all people with PCD and their families in Switzerland for participating in the survey and the CH-PCD. We are also grateful to the Swiss PCD support group that closely collaborates with us. We thank Eugenie Collaud (ISPM, University of Bern) for her contributions to the French translation of the questionnaire and Kristin Marie Bivens (ISPM, University of Bern) for her editorial assistance.

The current **Swiss PCD research group** includes (in alphabetical order): Juerg Barben (Kantonsspital St. Gallen), Sylvain Blanchon (University Hospital of Lausanne), Jean-Louis Blouin (University Hospitals of Geneva), Marina Bullo (Inselspital Bern), Carmen Casaulta (Inselspital Bern), Cristian Clarenbach (University Hospital of Zurich), Myrofora Goutaki (ISPM, University of Bern), Nicolas Gürtler (University Hospital of Basel, Switzerland), Beat Haenni (Institute of Anatomy Bern), Andreas Hector (University Children’s Hospital Zurich), Michael Hitzler (Children’s Hospital Lucerne), Andreas Jung (University Children’s Hospital Zurich), Lilian Junker (Hospital Thun), Elisabeth Kieninger (Inselspital Bern), Claudia E. Kuehni (ISPM, University of Bern), Yin Ting Lam (ISPM, University of Bern), Philipp Latzin (Inselspital Bern), Romain Lazor (University Hospital of Lausanne), Dagmar Lin (Bern University Hospital), Marco Lurà (Children’s Hospital Lucerne), Loretta Müller (Inselspital Bern), Eva Pedersen (ISPM, University of Bern), Nicolas Regamey (Children’s Hospital Lucerne), Isabelle Rochat (University Hospital of Lausanne), Daniel Schilter (Quartier Bleu Bern), Iris Schmid (Quartier Bleu Bern), Bernhard Schwizer (Quartier Bleu Bern), Andrea Stokes (Inselspital Bern), Daniel Trachsel (University Children’s Hospital Basel), Stefan A. Tschanz (Institute of Anatomy, University of Bern), Johannes Wildhaber (University of Fribourg), and Maura Zanolari (Hospital of Bellinzona).

## References

1. Fernandes AC, Bezerra OM. Nutrition therapy for chronic obstructive pulmonary disease and related nutritional complications. J Bras Pneumol 2006;32:461–71.

2. Cordova-Rivera L, Gibson PG, Gardiner PA, McDonald VM. Physical activity associates with disease characteristics of severe asthma, bronchiectasis and COPD. Respirology 2019;24:352–60.

3. Urquhart DS. Exercise testing in cystic fibrosis: why (and how)? J R Soc Med 2011;104 Suppl 1:S6–14.

4. Vogiatzis I, Zakynthinos G, Andrianopoulos V. Mechanisms of physical activity limitation in chronic lung diseases. Pulm Med 2012;2012:634761.

5. Barbato A, Frischer T, Kuehni CE, Snijders D, Azevedo I, Baktai G, Bartoloni L, Eber E, Escribano A, Haarman E, Hesselmar B, Hogg C, Jorissen M, Lucas J, Nielsen KG, O’Callaghan C, Omran H, Pohunek P, Strippoli MP, Bush A. Primary ciliary dyskinesia: a consensus statement on diagnostic and treatment approaches in children. Eur Respir J 2009;34:1264–76.

6. Goutaki M, Meier AB, Halbeisen FS, Lucas JS, Dell SD, Maurer E, Casaulta C, Jurca M, Spycher BD, Kuehni CE. Clinical manifestations in primary ciliary dyskinesia: systematic review and meta-analysis. Eur Respir J 2016;48:1081–95.

7. Valerio G, Giallauria F, Montella S, Vaino N, Vigorito C, Mirra V, Santamaria F. Cardiopulmonary assessment in primary ciliary dyskinesia. Eur J Clin Invest 2012;42:617–22.

8. Schofield LM, Duff A, Brennan C. Airway Clearance Techniques for Primary Ciliary Dyskinesia; is the Cystic Fibrosis literature portable? Paediatr Respir Rev 2018;25:73–7.

9. Kuehni CE, Goutaki M, Rubbo B, Lucas JS. Management of primary ciliary dyskinesia: current practice and future perspectives In: Chalmers JD, Polverino E, Aliberti S, eds. Bronchiectasis (ERS Monograph). 2018.

10. Kriemler S, Radtke T, Christen G, Kerstan-Huber M, Hebestreit H. Short-Term Effect of Different Physical Exercises and Physiotherapy Combinations on Sputum Expectoration, Oxygen Saturation, and Lung Function in Young Patients with Cystic Fibrosis. Lung 2016;194:659–64.

11. Nigro E, Polito R, Elce A, Signoriello G, Iacotucci P, Carnovale V, Gelzo M, Zarrilli F, Castaldo G, Daniele A. Physical Activity Regulates TNFα and IL-6 Expression to Counteract Inflammation in Cystic Fibrosis Patients. Int J Environ Res Public Health 2021;18.

12. Simsek S, Inal-Ince D, Cakmak A, Emiralioglu N, Calik-Kutukcu E, Saglam M, Vardar-Yagli N, Ozcelik HU, Sonbahar-Ulu H, Bozdemir-Ozel C, Kiper N, Arikan H. Reduced anaerobic and aerobic performance in children with primary ciliary dyskinesia. Eur J Pediatr 2018;177:765–73.

13. Madsen A, Green K, Buchvald F, Hanel B, Nielsen KG. Aerobic fitness in children and young adults with primary ciliary dyskinesia. PLoS One 2013;8:e71409.

14. Ring AM, Buchvald FF, Holgersen MG, Green K, Nielsen KG. Fitness and lung function in children with primary ciliary dyskinesia and cystic fibrosis. Respir Med 2018;139:79–85.

15. Firat M, Bosnak-Guclu M, Sismanlar-Eyuboglu T, Tana-Aslan A. Respiratory muscle strength, exercise capacity and physical activity in patients with primary ciliary dyskinesia: A cross-sectional study. Respir Med 2022;191:106719.

16. Goutaki M, Maurer E, Halbeisen FS, Amirav I, Barbato A, Behan L, Boon M, Casaulta C, Clement A, Crowley S, Haarman E, Hogg C, Karadag B, Koerner-Rettberg C, Leigh MW, Loebinger MR, Mazurek H, Morgan L, Nielsen KG, Omran H, Schwerk N, Scigliano S, Werner C, Yiallouros P, Zivkovic Z, Lucas JS, Kuehni CE. The international primary ciliary dyskinesia cohort (iPCD Cohort): methods and first results. Eur Respir J 2017;49.

17. Marino LV, Harris A, Johnstone C, Friend A, Newell C, Miles EA, Lucas JS, Calder PC, Walker WT. Characterising the nutritional status of children with primary ciliary dyskinesia. Clin Nutr 2019;38:2127–35.

18. Goutaki M, Halbeisen FS, Spycher BD, Maurer E, Belle F, Amirav I, Behan L, Boon M, Carr S, Casaulta C, Clement A, Crowley S, Dell S, Ferkol T, Haarman EG, Karadag B, Knowles M, Koerner-Rettberg C, Leigh MW, Loebinger MR, Mazurek H, Morgan L, Nielsen KG, Phillipsen M, Sagel SD, Santamaria F, Schwerk N, Yiallouros P, Lucas JS, Kuehni CE. Growth and nutritional status, and their association with lung function: a study from the international Primary Ciliary Dyskinesia Cohort. Eur Respir J 2017;50.

19. Goutaki M, Eich MO, Halbeisen FS, Barben J, Casaulta C, Clarenbach C, Hafen G, Latzin P, Regamey N, Lazor R, Tschanz S, Zanolari M, Maurer E, Kuehni CE. The Swiss Primary Ciliary Dyskinesia registry: objectives, methods and first results. Swiss Med Wkly 2019;149.

20. Lucas JS, Barbato A, Collins SA, Goutaki M, Behan L, Caudri D, Dell S, Eber E, Escudier E, Hirst RA, Hogg C, Jorissen M, Latzin P, Legendre M, Leigh MW, Midulla F, Nielsen KG, Omran H, Papon JF, Pohunek P, Redfern B, Rigau D, Rindlisbacher B, Santamaria F, Shoemark A, Snijders D, Tonia T, Titieni A, Walker WT, Werner C, Bush A, Kuehni CE. European Respiratory Society guidelines for the diagnosis of primary ciliary dyskinesia. Eur Respir J 2017;49.

21. Müller L, Savas ST, Tschanz SA, Stokes A, Escher A, Nussbaumer M, Bullo M, Kuehni CE, Blanchon S, Jung A, Regamey N, Haenni B, Schneiter M, Ingold J, Kieninger E, Casaulta C, Latzin P, On Behalf Of The Swiss Pcd Research G. A Comprehensive Approach for the Diagnosis of Primary Ciliary Dyskinesia-Experiences from the First 100 Patients of the PCD-UNIBE Diagnostic Center. Diagnostics (Basel) 2021;11.

22. von Elm E, Altman DG, Egger M, Pocock SJ, Gøtzsche PC, Vandenbroucke JP. The Strengthening the Reporting of Observational Studies in Epidemiology (STROBE) statement: guidelines for reporting observational studies. J Clin Epidemiol 2008;61:344–9.

23. Goutaki M, Hüsler L, Lam YT, Collaud E, Koppe H, Pedersen E, Kuehni C. Respiratory symptoms of Swiss people with Primary Ciliary Dyskinesia. European Respiratory Journal 2021;58:OA2956.

24. Goutaki M, Papon JF, Boon M, Casaulta C, Eber E, Escudier E, Halbeisen FS, Harris A, Hogg C, Honore I, Jung A, Karadag B, Koerner-Rettberg C, Legendre M, Maitre B, Nielsen KG, Rubbo B, Rumman N, Schofield L, Shoemark A, Thouvenin G, Willkins H, Lucas JS, Kuehni CE. Standardised clinical data from patients with primary ciliary dyskinesia: FOLLOW-PCD. ERJ Open Res 2020;6.

25. Physical Activity Guidelines for Americans, 2nd edition. Department of Health and Human Services, 2018. (Accessed 29.03.2022, at https://www.cdc.gov/physicalactivity/basics/index.htm.)

26. Garber CE, Blissmer B, Deschenes MR, Franklin BA, Lamonte MJ, Lee IM, Nieman DC, Swain DP. American College of Sports Medicine position stand. Quantity and quality of exercise for developing and maintaining cardiorespiratory, musculoskeletal, and neuromotor fitness in apparently healthy adults: guidance for prescribing exercise. Med Sci Sports Exerc 2011;43:1334–59.

27. Haskell WL, Lee IM, Pate RR, Powell KE, Blair SN, Franklin BA, Macera CA, Heath GW, Thompson PD, Bauman A. Physical activity and public health: updated recommendation for adults from the American College of Sports Medicine and the American Heart Association. Med Sci Sports Exerc 2007;39:1423–34.

28. Obesity: preventing and managing the global epidemic. Report of a WHO consultation. World Health Organ Tech Rep Ser 2000;894:i-xii, 1-253.

29. WHO Child Growth Standards based on length/height, weight and age. Acta Paediatr Suppl 2006;450:76–85.

30. Sport Schweiz 2020: Kinder-und Jugendbericht. 2021. (Accessed 29.03.2022, at https://www.admin.ch/gov/de/start/dokumentation/medienmitteilungen.msg-id-84993.html.)

31. Schindera C, Weiss A, Hagenbuch N, Otth M, Diesch T, von der Weid N, Kuehni CE. Physical activity and screen time in children who survived cancer: A report from the Swiss Childhood Cancer Survivor Study. Pediatr Blood Cancer 2020;67:e28046.

32. Rubbo B, Best S, Hirst RA, Shoemark A, Goggin P, Carr SB, Chetcuti P, Hogg C, Kenia P, Lucas JS, Moya E, Narayanan M, O’Callaghan C, Williamson M, Walker WT. Clinical features and management of children with primary ciliary dyskinesia in England. Arch Dis Child 2020;105:724–9.

33. Das Gewicht der Schweiz. Eine quantitative Synthesestudie zum Body Mass Index und Bauchumfang sowie den damit verbundenen Kofaktoren bei erwachsenen Männern und Frauen in der Schweiz. 2020. at https://gesundheitsfoerderung.ch/public-health/ernaehrung-und-bewegung-bei-kindern-und-jugendlichen/produkte-dienstleistungen/rechner-body-mass-index-bmi.html.)

34. Barak A, Wexler ID, Efrati O, Bentur L, Augarten A, Mussaffi H, Avital A, Rivlin J, Aviram M, Yahav Y, Kerem E. Trampoline use as physiotherapy for cystic fibrosis patients. Pediatr Pulmonol 2005;39:70–3.

35. Schofield LM, Horobin HE. Growing up with Primary Ciliary Dyskinesia in Bradford, UK: exploring patients experiences as a physiotherapist. Physiother Theory Pract 2014;30:157–64.

