## Supplement table S1 for "Physical activity, respiratory physiotherapy practices, and nutrition among people with primary ciliary dyskinesia in Switzerland"

**Supplemental Table S1:** Top 10 most common types of physical activity of people with PCD in Switzerland, overall and by age group. Some people reported multiple activities.

| | Total | Children <18 y | Adults ≥18 y |
|---|---|---|---|
| **Number of participants** | 74 | 24 | 50 |
| Training in gym | 17 | 0 | 17 |
| Walking | 10 | 0 | 10 |
| Cycling | 9 | 1 | 8 |
| Jogging | 4 | 0 | 4 |
| Ice hockey | 4 | 1 | 3 |
| Hiking | 2 | 0 | 2 |
| Horseback riding | 4 | 2 | 2 |
| Skiing/snowboarding | 3 | 1 | 2 |
| Badminton | 1 | 0 | 1 |
| Climbing | 1 | 0 | 1 |
